# Validation of reported risk factors for disease classification and prognosis in COVID-19: a descriptive and retrospective study

**DOI:** 10.1101/2020.04.05.20053769

**Authors:** Li Tan, Xia Kang, Xinran Ji, Qi Wang, Yongsheng Li, Qiongshu Wang, Hongming Miao

## Abstract

Risk indicators viral load (ORF1ab Ct), lymphocyte percentage (LYM%), C-reactive protein (CRP), interleukin-6 (IL-6), procalcitonin (PCT) and lactic acid (LA) in COVID-19 patients have been proposed in recent studies. However, the predictive effects of those indicators on disease classification and prognosis remains largely unknown. We dynamically measured those reported indicators in 132 cases of COVID-19 patients including the moderate-cured (moderated and cured), severe-cured (severe and cured) and critically ill (died). Our data showed that CRP, PCT, IL-6, LYM%, lactic acid and viral load could predict prognosis and guide classification of COVID-19 patients in different degrees. CRP, IL-6 and LYM% were more effective than other three factors in predicting prognosis. For disease classification, CRP and LYM% were sensitive in identifying the types between critically ill and severe (or moderate). Notably, among the investigated factors, LYM% was the only one that could distinguish between the severe and moderate types. Collectively, we concluded that LYM% was the most sensitive and reliable predictor for disease typing and prognosis. During the COVID-19 pandemic, the precise classification and prognosis prediction are critical for saving the insufficient medical resources, stratified treatment and improving the survival rate of critically ill patients. We recommend that LYM% be used independently or in combination with other indicators in the management of COVID-19.

## Introduction

Coronavirus disease-19 (COVID-19) is an acute respiratory infective disease caused by severe acute respiratory syndrome coronavirus 2 (SARS-CoV-2)[1-2]. This pandemic has been spreading worldwide rapidly since March 2020. It was reported that more than 500,000 individuals had been diagnosed and this disease caused over 20,000 deaths at late of March 2020. The rapidly increased cases lead to overload of public healthcare sources and a run on medical facilities[3]. The severity of patients with COVID-19 can be divided into 4 levels, i.e. mild, moderate, severe and critical according to the New Coronavirus Pneumonia Diagnosis Program (5th edition) published by the National Health Commission of China[4]. Current experience reveals that most of infected people (approx. over 80%) are not severe and can get self-healing, only a small part of cases should be carefully treated and hospitalized[5-8]. However, the mortality among the severe, especially the critical patients, is quite high, thus it is critical to screen out reliable predictors for severity of patients to get better outcomes and to save medical sources.

Several factors including viral load[9-10], lymphocytes[11-12], C-reactive protein (CRP)[8, 13], interleukin-6 (IL-6)[14-15], procalcitonin (PCT)[16-17] and lactic acid[7, 17] have been identified as warning indicators of prognosis in COVID-19 patients in recent studies from different cohorts of patients. However, it is still elusive that which of these factors are the most effective and reliable indicators for predicting the prognosis of COVID-19 in the early stage.

Besides, the indicators for disease classification were also urgently needed. In the present study, basing on the clinical information of 132 patients with COVID-19, we will compare and validate the predictive power of several reported risk factors of prognosis in COVID-19 in a descriptive manner.

## Methods

### Patient information

All cases were taken from the General Hospital of Central Theater Command (Wuhan, Hubei province, People’s Republic of China). This study was approved by the Ethics Committee of the hospital. All subjects signed informed consent forms at admission to hospital. A total of 132 patients hospitalized between January 14, 2020 and March 14, 2020 were investigated, of which 96 cured cases were moderate type (moderate-cured), 21 cured cases were severe type (severe-cured) and 15 dead cases were critically ill type (critically ill-died). The gender and age information of those patients were presented in Table S1 and S2. All patients were confirmed by viral detections of SARS-CoV-2 via oropharyngeal swabs using quantitative RT-PCR for ORF1ab[18], which ruled out infection by other respiratory viruses such as influenza virus A, influenza virus B, coxsackie virus, respiratory syncytial virus, parainfluenza virus and enterovirus.

### Disease classification

All cases were diagnosed and classified according to the New Coronavirus Pneumonia Diagnosis Program (5th edition) published by the National Health Commission of China[4]. Clinical manifestations consist of four categories, mild, moderate, severe and critically ill. The mild clinical symptom were mild with no pulmonary inflammation on imaging. The moderate is the overwhelming majority, showing symptoms of respiratory infections such as fever, cough, and sputum, and pulmonary inflammation on imaging; when symptoms of dyspnea appear, including any of the following: shortness of breath, RR ≥ 30bpm, blood oxygen saturation ≤ 93% (at rest), PaO2 / FiO2 ≤ 300 mmHg, or pulmonary inflammation that progresses significantly within 24 to 48 hours> 50%, it was classified as severe; respiratory failure, shock, and organ failures that require intensive care were critically ill. Among them, mild patients were not admitted in this designated hospital.

### Data collection

In this study, the basic information, complete blood count, serum biochemical test, inflammatory indicators, viral load and disease outcome of all included patients were collected dynamically.

### Statistical methods

In this study, GraphPad 8.01 software was used for data statistics and mapping. Data were expressed as the means±s.e.ms. and were analyzed using two-tailed unpaired Student’s *t*-test. For each parameter of all data presented, * indicates P< 0.05, ** indicates P<0.01 and *** indicates P<0.005..

## Results

### Validation of the risk factors for prognosis in COVID-19

To investigate the mortality-associated risk factors, COVID-19 patients were classified into cured and died groups (Table S1). The serum levels of CRP, PCT, IL-6, percentage of lymphocytes (LYM%), lactic acid and Ct values of viral tests were measured dynamically. The levels of CRP in peripheral blood increased quickly after the symptom onset and further increased to the end around 4 weeks later in the dead cases, while the levels of CRP in survivors fluctuated very little (Fig. 1A). Similar trend was obtained for the PCT level, although it just began to increase 25 days after disease onset (Fig. 1B). The blood IL-6 levels in dead cases increased rapidly in the early phase and rose to an extraordinary level before death (Fig. 1C). In contrast, after the symptom onset, the LYM% in dead cases deceased quickly and sustained in a low level (Fig. 1D). For the levels of blood lactic acid, there was a narrow time window in which we could distinguish the died and cured groups (Fig. 1E). From 8 days after disease onset, the pre-died patients had an obviously higher levels of viral load indicated by ORF1ab Ct values than the survivors did (Fig. 1F). Taken together, indicators CRP, PCT IL-6, LYM% and viral load could predict the mortality risk in different degrees. IL-6, LYM% and CRP are sensitive indicators because obvious changes of those factors in the early phase (2 or 4 days after disease onset) were identified between the cured and died groups (Fig. 1G). In addition, those three indicators had a long time window in distinguishing cured and died cases, which could ensure the accuracy of the forecast(Fig. 1G).

**Fig. 1.**
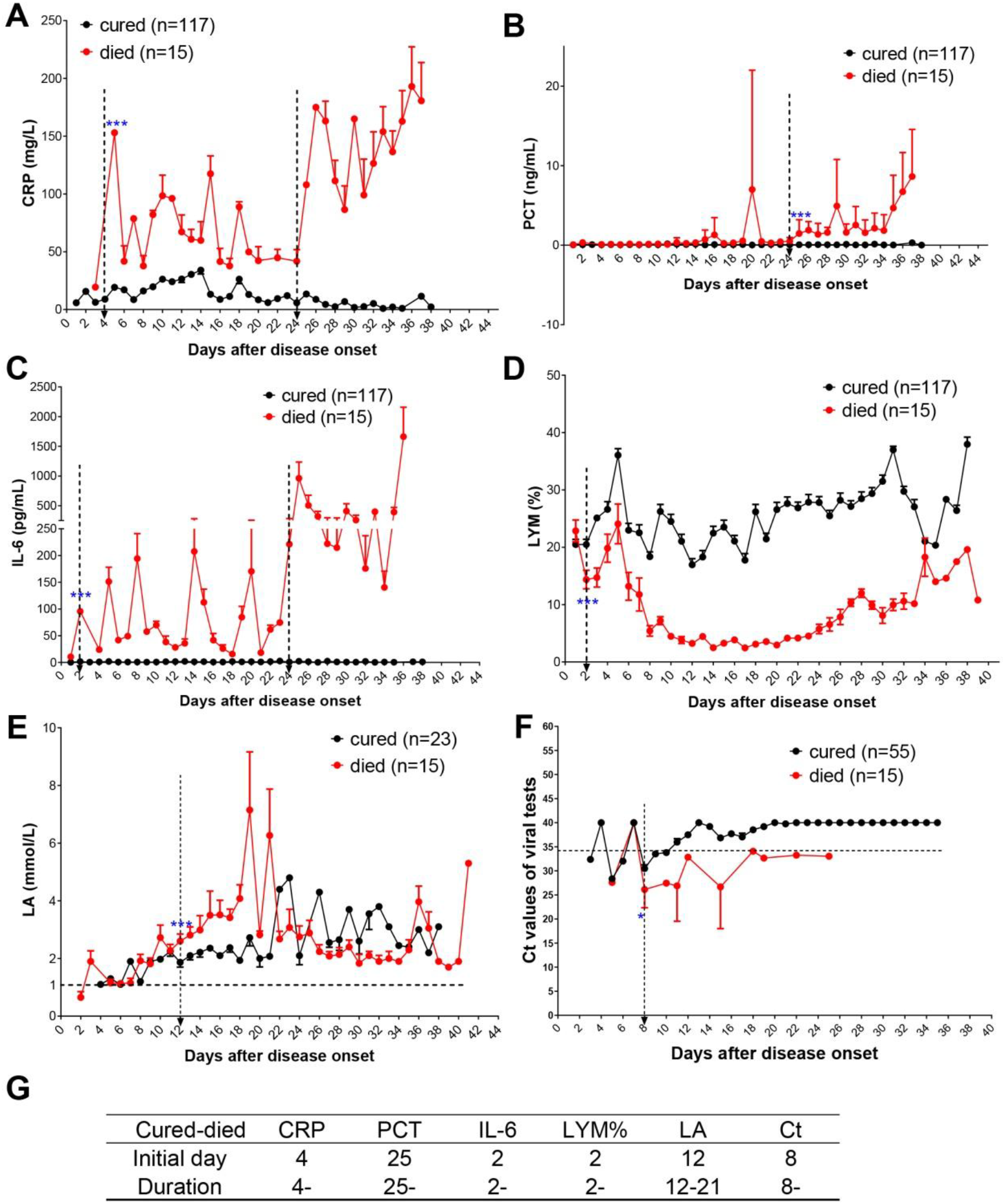
Dynamic changes of indicators for prognosis of COVID-19 patients. (**A-D)** Dynamic changes of CRP(**A**), PCT (**B**), IL-6 (**C**) and LYM% (**D**) in the peripheral blood in the cured (n=117) or died (n=15) patients with COVID-19. (E) Dynamic changes of LA levels in the blood of cured (n=23) or died (n=15) patients with COVID-19. **(F)** Dynamic changes of ORF1ab Ct values in viral tests with quantitative RT-PCR in cured (n=55) or died (n=15) patients with COVID-19. **(G)** Time windows of indicators for predicting prognosis. Initial day indicates the first time with significant differences between two groups after disease onset. Duration indicates the persistent difference for more than 7 days. The duration less than 7 days was not counted. The dotted arrow indicates an important cutoff of timeline. Data showed means ± s.e.ms. (*P<0.05 and ***P<0.005; Student’s *t* test) CRP, C-reactive protein; PCT, procalcitonin; LYM%, lymphocyte percentage; LA, lactic acid;

### Validating the indicators for classifying the critically ill and severe types in COVID-19

The initiation and duration of difference in some indicator between two groups are the key to judge the disease types. In distinguishing between the critically ill and severe groups, the initial day of difference for indicators CRP, PCT, IL-6, LYM%, lactic acid and ORF1ab Ct was 4, 25, 2, 3, 12 and 11, respectively (Fig. 2A-G). The duration of difference for indicators CRP, PCT, IL-6, LYM%, lactic acid and ORF1ab Ct was 4-, 25-, 22-, 8-, 12-24 and 19-, respectively (Fig. 2A-G). These results indicated that CRP, IL-6 and LYM% were more sensitive and reliable than other three indicators for disease typing between the critically ill and severe.

**Fig. 2.**
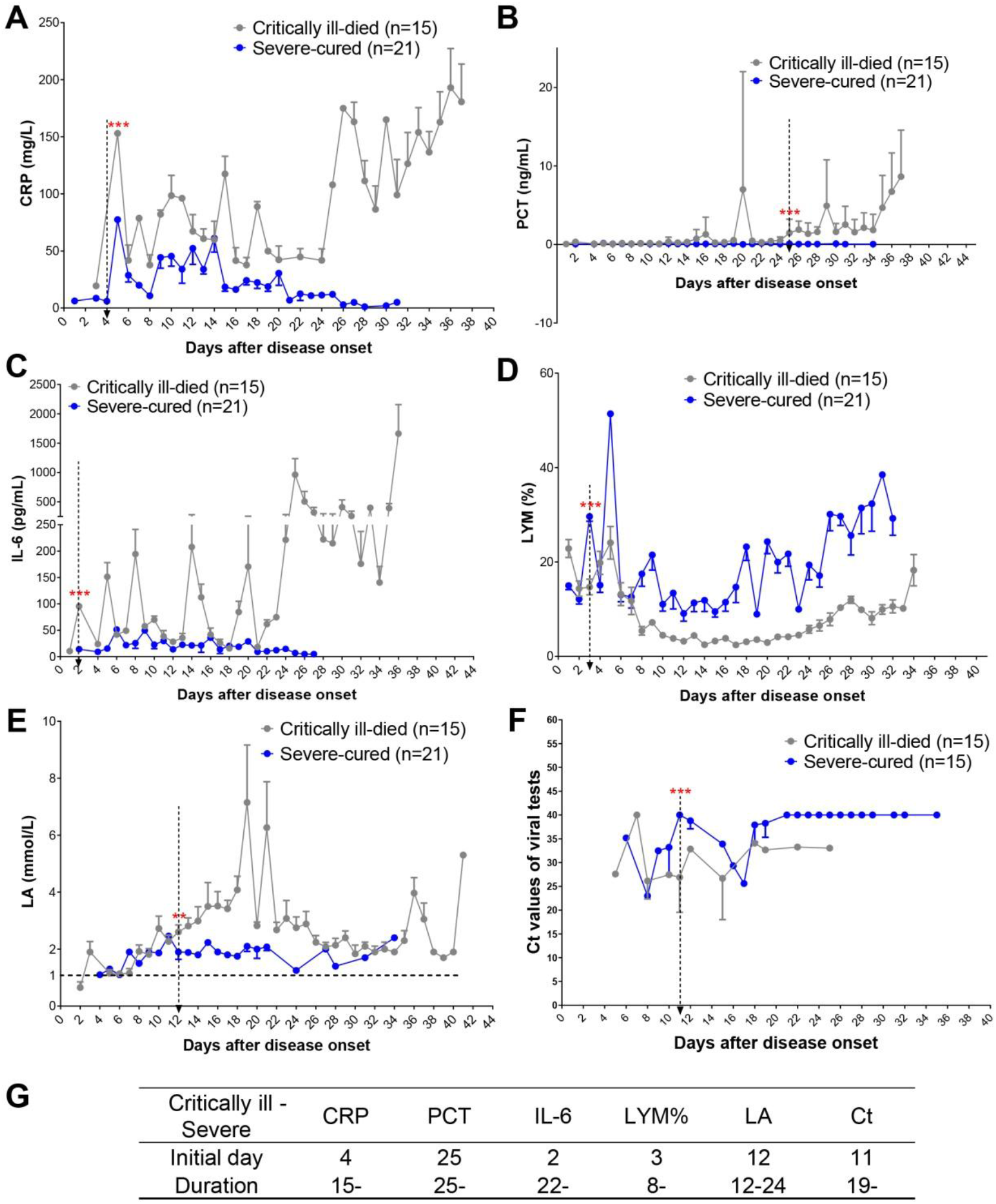
Dynamic changes of indicators in COVID-19 patients with critically ill or severe types. (**A-E)** Dynamic alterations of CRP(**A**), PCT (**B**), IL-6 (**C**), LYM% (**D**) and LA in the peripheral blood of the severe-cured (n=21) or critically ill (n=15) patients with COVID-19. **(F)** Dynamic changes of ORF1ab Ct values in severe-cured (n=15) or criticallyill (n=15) patients with COVID-19. **(G)** Time windows of indicators for disease classification between critically ill and severe types. Data showed means ± s.e.ms. (**P<0.01 and ***P<0.005; Student’s *t* test)

### Validating the indicators for classifying the critically ill and moderate COVID-19 patients

To study the indicators classifying the critically ill and moderate patients, those two types of patients were enrolled. The initial day of difference for indicators CRP, PCT, IL-6, LYM% and ORF1ab Ct was 4, 25, 2, 2 and 8, respectively (Fig. 3A-F). The duration of difference for indicators CRP, PCT, IL-6, LYM% and ORF1ab Ct was 4-, 25-, 22-, 2- and 8-, respectively (Fig. 3A-F). Taken together, CRP and LYM% were more effective in typing the critically ill and moderate cases.

**Fig. 3.**
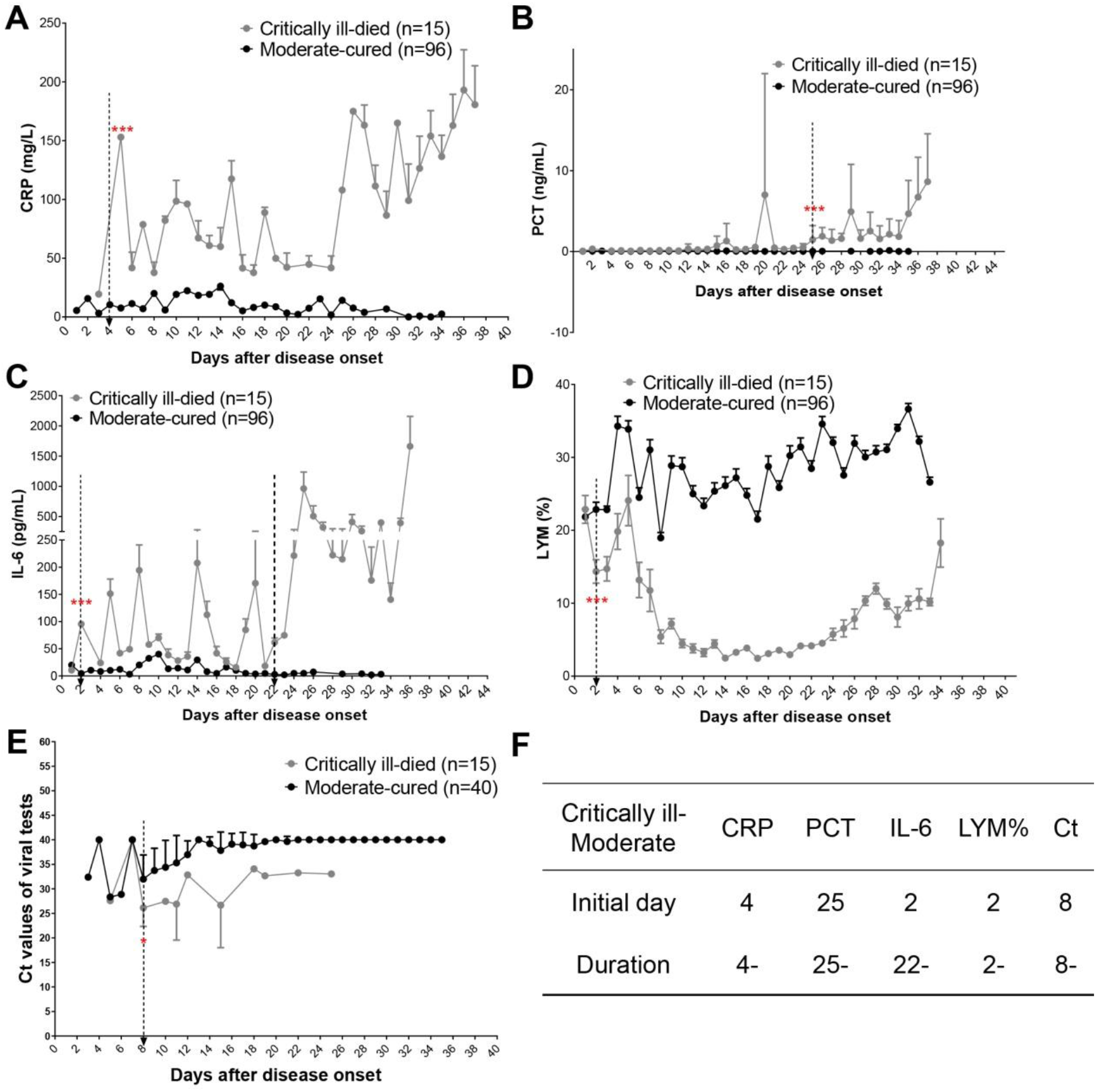
Dynamic alterations of indicators in COVID-19 patients with critically ill or moderate types. (**A-D)** Dynamic alterations of CRP(**A**), PCT (**B**), IL-6 (**C**) and LYM% (**D**) in the peripheral blood of the critically ill (n=15) or moderate (n=96) patients with COVID-19. **(E)** Dynamic changes of ORF1ab Ct values in the critically ill (n=15) or moderate (n=40) patients with COVID-19. **(F)** Time windows of indicators for disease typing between critically ill and moderate types. Data showed means ± s.e.ms. (*P<0.05 and ***P<0.005; Student’s *t* test)

### Validating the indicators for classifying the severe and moderate COVID-19 patients

For the indicators of the severe and moderate types, the initial day of difference for indicators CRP, IL-6, LYM% and ORF1ab Ct was 5, 6, 1 and 8, respectively. In contrast, there were totally no difference of PCT levels in both groups (Fig. 4A-F). Noteworthily, the duration of difference for CRP and LYM% was 9-20 and 9-26, respectively, while PCT, IL-6 and ORF1ab Ct had no continuous time window for differentiation (Fig. 4A-F). These results indicated that LYM% was the only indicator among those 5 factors for the typing of severe and moderate patients.

**Fig. 4.**
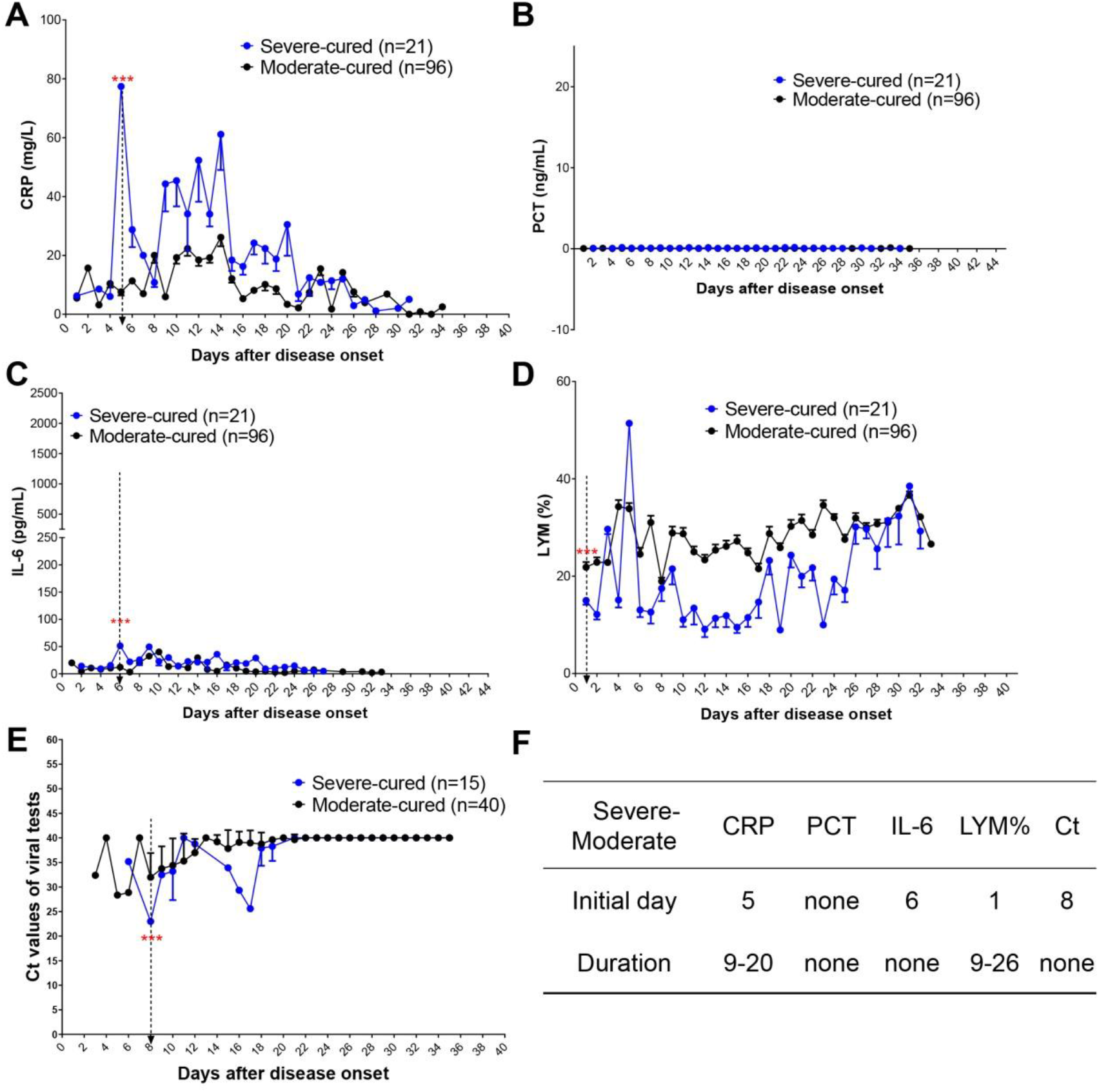
Dynamic alterations of indicators in COVID-19 patients with severe or moderate types. **(A-D)** Dynamic alterations of CRP(**A**), PCT (**B**), IL-6 (**C**) and LYM% (**D**) in the peripheral blood of the severe (n=21) or moderate (n=96) patients with COVID-19. **(E)** Dynamic changes of ORF1ab Ct values in the severe (n=15) or moderate (n=40) patients with COVID-19. **(F)** Time windows of indicators for disease typing between severe and moderate types. Data showed means ± s.e.ms. (***P<0.005; Student’s *t* test)

## Discussion

Given the worldwide prevalence of COVID-19, disease typing and prognostic indicators are of great significance in guiding classified treatment, preventing medical runs and saving critical patients. In this study, we selected several reported risk factors for prognosis of COVID-19 patients and further validated their predictive roles in disease outcome and classification. This is the first study reporting CRP, PCT, IL-6, lactic acid, ORF1ab Ct and LYM% as indicators for different types of patients with COVID-19.

We validated that the LYM% could be a permissible predictor to identify critically ill, severe and moderate cases. Previous studies also supported the conclusion that lymphocyte count and function were closely related to the disease status of COVID-19[11, 19]. Patients with insufficient immunity, including the elders and people with immunodeficiency, always presented a lower level of lymphocytes and a worse prognosis after infected with SARS-CoV-2[7, 20]. In addition, the relative stability of LYM% in each patients and less overlap among groups with different severity during the progression of COVID-19 made this indicator a potential good predictor.

Notably, as we described above, the serum levels of IL-6 and CRP presented with a zigzag curve during the whole course of disease in critically ill patients. We speculated that this obvious change may be caused by the medical intervention. Interestingly, the changes of serum levels of these two markers in moderate and sever patients were much more stable and lower than those in critical patients. These results indicated that refractory inflammatory reaction can be identified in early stage of this disease and predict a poor outcome. Patients with this feature should be brought to the forefront in early stage. In clinical practice, the curve of inflammatory indicators should be draw dynamically, like body temperature.

The higher levels of inflammatory indicators and the lower level of LYM% leads to another interesting question: whether the “storm” of inflammatory cytokines is due to the impaired lymphocytes, such as T cells. As we know, some of T cells, i.e. Treg, are responsible for inflammatory regulation[21-22]. The impaired function of Treg cells may contribute to the uncontrolled inflammation in critical patients. It should be noted that in the late stage of dead cases, all the levels of proinflammatory indicators, LYM% and lactic acid had an acute increase (Fig. 1A-E). This phenomenon indicated an absolute disorder of inflammation, immunity and metabolism in those dying patients.

Persistently low levels of Ct values in viral tests indicated sustained high levels of viral load in the critically ill patients[10]. However, it should be noted that there were no obvious differences in the duration of viral shedding between the severe and non-severe patients, which was consistent with a previous study[7]. Recently, we reported a moderate patient with long duration of viral shedding for 49 days. This phenomenon was in line with the asymptomatic infection, which was reported in some population[23-24].

## Conclusion

In the present study, basing on the information of 132 patients with COVID-19, we analyzed the predictive power of several reported indicators for disease severity and prognosis in a descriptive manner. We found that CRP, PCT, IL-6, LYM%, lactic acid and viral load could predict prognosis and guide classification of COVID-19 patients in different degrees. CRP, IL-6 and LYM% were more effective than other three factors in predicting prognosis. CRP and LYM% were sensitive in identifying the types between critically ill and severe (or moderate). Notably, among the investigated factors, LYM% was the only one that could distinguish between the severe and moderate types. Collectively, we concluded that LYM% was the most sensitive and reliable predictor for disease typing and prognosis. We recommend that LYM% be used independently or in combination with other indicators in the management of COVID-19.

## Data Availability

All the data were included in the paper or supplementary materials.The raw data could be provided by the corresponding authors upon reasonable request.

## Acknowledgements

We thank all the medical and scientific workers for the efforts in fighting against SARS-CoV-2. This study was supported in part by award numbers 81872028 and 81672693 (H.M.) from the National Natural Science Foundation of China, cstc2017jcyjBX0071 (H.M.) from the Foundation and Frontier Research Project of Chongqing and T04010019 (H.M.) from the Chongqing Youth Top Talent Project.

## Contributions

Li Tan, Qiongshu Wang and Qi Wang were responsible for data collection. Xia Kang, Xinran Ji and Yongsheng Li analyzed the data and contributed to discussion. Hongming Miao was responsible for conceptual design, manuscript writing and submission. All authors read the manuscript and approved the submission.

## Supplementary tables

**Table S1.** Gender and age information of the cured or died COVID-19 patients

|  | Gender(Male/Female) | Age(Male/Female) |
| --- | --- | --- |
| Cured (n=117) | 46 (39%)/71 (61%) | 53 (54/52) |
| Died (n=15) | 13 (87%)/2 (13%) | 75 (79/45) |

**Table S2.** Gender and age information of the COVID-19 patients with different disease classification

|  | Gender(Male/Female) | Age(Male/Female) |
| --- | --- | --- |
| Critically ill-died (n=15) | 13 (87%)/2 (13%) | 75 (79/45) |
| Severe-cured (n=21) | 11 (52%)/10 (48%) | 51 (49/53) |
| Moderate-cured (n=96) | 36 (38%)/60 (62%) | 53 (56/51) |

